# Comparison of two highly-effective mRNA vaccines for COVID-19 during periods of Alpha and Delta variant prevalence

**DOI:** 10.1101/2021.08.06.21261707

**Authors:** Arjun Puranik, Patrick J. Lenehan, Eli Silvert, Michiel J.M. Niesen, Juan Corchado-Garcia, John C. O’Horo, Abinash Virk, Melanie D. Swift, John Halamka, Andrew D. Badley, A.J. Venkatakrishnan, Venky Soundararajan

**Author notes:** Correspondence to: Venky Soundararajan. These authors contributed equally (Arjun Puranik, Patrick J. Lenehan).

## Abstract

Although clinical trials and real-world studies have affirmed the effectiveness and safety of the FDA-authorized COVID-19 vaccines, reports of breakthrough infections and persistent emergence of new variants highlight the need to vigilantly monitor the effectiveness of these vaccines. Here we compare the effectiveness of two full-length Spike protein-encoding mRNA vaccines from Moderna (mRNA-1273) and Pfizer/BioNTech (BNT162b2) in the Mayo Clinic Health System over time from January to July 2021, during which either the Alpha or Delta variant was highly prevalent. We defined cohorts of vaccinated and unvaccinated individuals from Minnesota (n = 25,589 each) matched on age, sex, race, history of prior SARS-CoV-2 PCR testing, and date of full vaccination. Both vaccines were highly effective during this study period against SARS-CoV-2 infection (mRNA-1273: 86%, 95%CI: 81-90.6%; BNT162b2: 76%, 95%CI: 69-81%) and COVID-19 associated hospitalization (mRNA-1273: 91.6%, 95% CI: 81-97%; BNT162b2: 85%, 95% CI: 73-93%). In July, vaccine effectiveness against hospitalization has remained high (mRNA-1273: 81%, 95% CI: 33-96.3%; BNT162b2: 75%, 95% CI: 24-93.9%), but effectiveness against infection was lower for both vaccines (mRNA-1273: 76%, 95% CI: 58-87%; BNT162b2: 42%, 95% CI: 13-62%), with a more pronounced reduction for BNT162b2. Notably, the Delta variant prevalence in Minnesota increased from 0.7% in May to over 70% in July whereas the Alpha variant prevalence decreased from 85% to 13% over the same time period. Comparing rates of infection between matched individuals fully vaccinated with mRNA-1273 versus BNT162b2 across Mayo Clinic Health System sites in multiple states (Minnesota, Wisconsin, Arizona, Florida, and Iowa), mRNA-1273 conferred a two-fold risk reduction against breakthrough infection compared to BNT162b2 (IRR = 0.50, 95% CI: 0.39-0.64). In Florida, which is currently experiencing its largest COVID-19 surge to date, the risk of infection in July after full vaccination with mRNA-1273 was about 60% lower than after full vaccination with BNT162b2 (IRR: 0.39, 95% CI: 0.24-0.62). Our observational study highlights that while both mRNA COVID-19 vaccines strongly protect against infection and severe disease, further evaluation of mechanisms underlying differences in their effectiveness such as dosing regimens and vaccine composition are warranted.

## Introduction

The SARS-CoV-2 virus has infected over 190 million individuals, leading to over 4 million deaths attributed to COVID-19.^1^ To curb the spread of SARS-CoV-2, mass global vaccination efforts have been initiated, with 3.9 billion vaccine doses administered to date.^2^ Controlled clinical trials and real-world clinical studies have provided clear evidence of the effectiveness of FDA-authorized COVID-19 vaccines. In clinical trials, BNT162b2, an mRNA vaccine developed by Pfizer/BioNTech, showed 95.0% efficacy (95% CI: 90.3-97.6%) in preventing symptomatic COVID-19 with onset seven or more days after the second dose.^3^ mRNA-1273, an mRNA vaccine developed by Moderna, showed 94.1% efficacy (95% CI: 89.3-96.8%) in preventing symptomatic infection with onset at least 14 days after the second dose.^4^ Additional real-world retrospective studies in major health systems in the United States and Israel further support the effectiveness and safety of these vaccines.^5–7^

However, only about 50% of the United States population is fully vaccinated as of July 2021, with an even lower fraction fully vaccinated across the globe.^2^ Further, there have been reports of reduced vaccine effectiveness against emerging variants and local increases in COVID-19 cases despite mass vaccination, raising questions about the potential need to administer vaccine booster doses and to develop variant-targeted vaccines in the future.^8–10^ This evolving state of affairs highlights the need to assess the durability and comparative effectiveness of the FDA-authorized vaccines. Here, we address this need by comparing the rates of SARS-CoV-2 infection and COVID-19 associated complications between demographically and geographically matched individuals who were vaccinated with mRNA-1273 versus BNT162b2 in the multi-state Mayo Clinic Health System.

## Methods

### Study design and population

This is a retrospective study of individuals who underwent SARS-CoV-2 polymerase chain reaction (PCR) testing at the Mayo Clinic and hospitals affiliated with the Mayo Clinic Health System (Arizona, Florida, Iowa, Minnesota, and Wisconsin). Overall, there were 645,109 individuals with at least one SARS-CoV-2 PCR test. We included individuals who met the following criteria: (i) age greater than or equal to 18 years; (ii) received at least one dose of BNT162b2 or mRNA-1273 after December 1, 2020 and on or before July 29, 2021; (iii) did not have any positive SARS-CoV-2 PCR tests prior to their first vaccine dose; and (iv) did not receive a mismatched series of COVID-19 vaccines (e.g., did not receive doses from more than one manufacturer). There were 119,463 individuals who met these criteria for BNT162b2 and 60,083 individuals who met these criteria for mRNA-1273.

### Defining matched cohorts of vaccinated and unvaccinated individuals

To determine both absolute and relative vaccine effectiveness, we used an exact matching procedure to construct cohorts of demographically and clinically similar individuals who were unvaccinated, vaccinated with mRNA-1273, or vaccinated with BNT162b2. Specifically, we attempted to identify “matched triples” as a set of three individuals (one unvaccinated, one who received mRNA-1273, and one who received BNT162b2) who were matched on the following criteria:

1. *Age (bucketed match)*. All individuals were classified into one of the following age buckets: 18-24, 25-34, 35-44, 45-54, 55-64, 65-74, 75-84, or 85+ years. For a given matched triple, all three individuals must be in the same bucket.
2. *Sex (exact match)*
3. *Race (exact match)*
4. *Ethnicity (exact match)*
5. *State of residence (exact match)*. This match helps to control for variability in (i) the vaccine rollout process (i.e., timeline and definition of eligible populations), (ii) community transmission patterns, and (iii) the dynamic landscape of SARS-CoV-2 variant prevalence between states.
6. *SARS-CoV-2 PCR testing history (bucketed match)*. All individuals were classified as having 0, 1, or multiple SARS-CoV-2 PCR tests (i) until December 1, 2020 and (ii) between December 1, 2020 and the date of their first vaccine dose. To be considered as a possible match, individuals had to match both of these bucketed classifications. This is intended to control for access to and/or likelihood of seeking out COVID-19 testing, as well as the baseline risk of exposure to SARS-CoV-2.
7. *Date of vaccination (bucketed)*. For a given individual in the mRNA-1273 cohort who received their first vaccine dose on a given date, only individuals in the BNT162b2 cohort who were vaccinated on the same date or within two weeks after that date were considered for matching. This match helps to ensure that matched individuals reach their date of full vaccination (14 days after the second dose) on approximately the same date.

This matching procedure yielded 43,895 matched triples. Of the 43,895 mRNA-1273-vaccinated individuals, 35,902 were fully vaccinated. Of the 43,895 BNT162b2-vaccinated individuals, 37,573 were fully vaccinated (**Table S1**). Unvaccinated individuals were assigned dates of hypothetical vaccination based on the actual vaccination dates of their matched partners. Specifically, the hypothetical first vaccination date for a given individual was defined as the date exactly halfway between actual first vaccination dates for the matched mRNA-1273- and BNT162b2-vaccinated individuals (rounding down when there is an odd number of days between). Similarly, when applicable, the hypothetical second vaccination date for a given individual was defined as the date exactly halfway between actual second vaccination dates for the matched mRNA-1273- and BNT162b2-vaccinated individuals. For cases in which only one of the vaccinated individuals had received a second dose, the hypothetical second vaccination date was taken as the exact same date of the single actual second vaccination date. When neither of the vaccinated individuals received a second dose, the hypothetical second vaccination date was taken to be the date exactly halfway between the suggested second dose dates of the vaccinated individuals, as long as this calculated date was prior to the study end date (July 30, 2021); if it was after the end of the study period, then no hypothetical second dose date was assigned. The distribution of actual or hypothetical second dose dates for the three cohorts is shown in **Figure S1**.

### Defining clinical outcomes of interest

To perform overall and comparative analyses of vaccine effectiveness, the following outcomes were assessed for each cohort:

1. *SARS-CoV-2 infection*: at least one positive SARS-CoV-2 PCR test. The date of infection was taken as the date of the first positive test.
2. *COVID-19 associated hospitalization*: admission to the hospital occurring within 21 days after SARS-CoV-2 infection.
3. *COVID-19 associated ICU admission*: admission to the intensive care unit (ICU) occurring within 21 days after SARS-CoV-2 infection.
4. *COVID-19 associated mortality*: death occurring within 28 days after SARS-CoV-2 infection.
5. *Breakthrough infection*: a SARS-CoV-2 infection occurring after full vaccination (i.e., at least 14 days after the second dose of mRNA-1273 or BNT162b2).

For each outcome, incidence rates (IRs) in cases per 1000 person-days were calculated for each cohort by dividing the number of cases (i.e. people experiencing the given outcome) by the total number of at-risk person-days and multiplying by 1000. For analyses of baseline risk, we considered only events occurring during the week after the first vaccine dose, during which vaccination is not yet expected to confer protection against infection.^3,4^ Here, each individual contributed at-risk person days from the date of their first actual or hypothetical vaccine dose until (i) they were infected with SARS-CoV-2, (ii) they died, (iii) the end of the study period (July 30, 2021), or (iv) seven days after their first dose (whichever came first). For analyses of breakthrough risks, we considered only events occurring after full vaccination was achieved (i.e., 14 days after the second actual or hypothetical vaccine dose).^11^ Here, each individual contributed at-risk person days from 14 days after their second dose until (i) they were infected with SARS-CoV-2, (ii) they died, or (iii) the end of the study period on July 30, 2021 (whichever came first).

To perform comparative analyses of breakthrough infection severity, the following outcomes were assessed for each cohort:

1. *21-day hospitalization*: the number of patients who were admitted to the hospital within 21 days of breakthrough infection diagnosis divided by the total number of patients with at least 21 days of follow-up after such diagnosis.
2. *21-day ICU admission*: the number of patients who were admitted to the ICU within 21 days of breakthrough infection diagnosis divided by the total number of patients with at least 21 days of follow-up after such diagnosis.
3. *28-day mortality*: the number of patients who died within 28 days of breakthrough infection diagnosis divided by the total number of patients with at least 28 days of follow-up after such diagnosis.

### Estimating vaccine effectiveness against SARS-CoV-2 infection and severe COVID-19

To estimate vaccine effectiveness, we compared the incidence rates of a given outcome (e.g., positive SARS-CoV-2 testing) between vaccinated and unvaccinated individuals. For this analysis, it is particularly important that individuals in the unvaccinated cohort are truly unvaccinated, which can be challenging to confirm because many people have been vaccinated outside of their primary care setting. There are differences in the methods and frequency of linking vaccination registries to the Mayo Clinic EHR between states, with Minnesota offering the distinct advantage of having automated biweekly syncing in place for its set of primary care patients. Thus, for all estimates of effectiveness (i.e., comparisons of vaccinated to unvaccinated individuals), we only considered the 25,859 matched triples of individuals from Minnesota. This cohort is summarized in **Table 1**.

**Table 1.** Clinical characteristics of 1-to-1 matched mRNA-1273-vaccinated versus BNT162b2-vaccinated versus unvaccinated cohorts in Minnesota. Covariates for matching include demographics (age, sex, race, ethnicity), state of residence, date of vaccination, and number of prior SARS-CoV-2 PCR tests. Covariate statistics are also shown for the sub-cohorts of each cohort which are counted in the full vaccination effectiveness analysis (i.e., patients who have received two doses of the given vaccine and had at least 14 days of follow-up after the second dose, without a positive PCR test at any date prior to 14 days following the second dose).

| Clinical covariate | Matched mRNA-1273, BNT162b2, unvaccinated *exactly* matched cohorts | Matched mRNA-1273 vaccinated cohort, fully vaccinated patients | Matched BNT162b2 vaccinated cohort, fully vaccinated patients | Matched unvaccinated cohort, 14+ days after hypothetical second dose | Maximum absolute SMD over the 3 pairs |
| --- | --- | --- | --- | --- | --- |
| Total number of individuals | 25,869 each | 21,179 | 22,064 | 24,990 |  |
| Age groups in years |  |  |  |  |  |
| - 18-24 | 1,668 (6.4%) | 1,214 (5.7%) | 1,301 (5.9%) | 1,601 (6.4%) | 0.03 |
| - 25-34 | 2,447 (9.5%) | 1,904 (9.0%) | 1,990 (9.0%) | 2,350 (9.4%) | 0.01 |
| - 35-44 | 2,931 (11.3%) | 2,338 (11.0%) | 2,352 (10.7%) | 2,833 (11.3%) | 0.02 |
| - 45-54 | 3,206 (12.4%) | 2,596 (12.3%) | 2,617 (11.9%) | 3,086 (12.3%) | 0.01 |
| - 55-64 | 5,104 (19.7%) | 4,283 (20.2%) | 4,265 (19.3%) | 4,942 (19.8%) | 0.02 |
| - 65-74 | 6,865 (26.5%) | 5,821 (27.5%) | 6,181 (28.0%) | 6,706 (26.8%) | 0.03 |
| - 75-84 | 2,440 (9.4%) | 2,038 (9.6%) | 2,240 (10.2%) | 2,348 (9.4%) | 0.03 |
| - 85+ | 1,208 (4.7%) | 985 (4.7%) | 1,118 (5.1%) | 1,124 (4.5%) | 0.03 |
| Sex |  |  |  |  |  |
| - Female | 14,613 (56.5%) | 11,973 (56.5%) | 12,515 (56.7%) | 14,137 (56.6%) | 0.00 |
| - Male | 11,256 (43.5%) | 9,206 (43.5%) | 9,549 (43.3%) | 10,853 (43.4%) | 0.00 |
| Race |  |  |  |  |  |
| - Asian | 307 (1.2%) | 255 (1.2%) | 276 (1.3%) | 299 (1.2%) | 0.00 |
| - Black / African American | 364 (1.4%) | 281 (1.3%) | 294 (1.3%) | 346 (1.4%) | 0.00 |
| - Native American | 15 (0.1%) | 11 (0.1%) | 10 (0.0%) | 15 (0.1%) | 0.01 |
| - White / Caucasian | 24,556 (94.9%) | 20,154 (95.2%) | 20,991 (95.1%) | 23,737 (95.0%) | 0.01 |
| - Other | 317 (1.2%) | 246 (1.2%) | 252 (1.1%) | 298 (1.2%) | 0.00 |
| - Unknown | 310 (1.2%) | 232 (1.1%) | 241 (1.1%) | 295 (1.2%) | 0.01 |
| Ethnicity |  |  |  |  |  |
| - Hispanic or Latino | 526 (2.0%) | 404 (1.9%) | 417 (1.9%) | 501 (2.0%) | 0.01 |
| - Not Hispanic or Latino | 24,822 (96.0%) | 20,378 (96.2%) | 21,225 (96.2%) | 23,996 (96.0%) | 0.01 |
| - Unknown | 521 (2.0%) | 397 (1.9%) | 422 (1.9%) | 493 (2.0%) | 0.01 |
| Number of PCR tests taken prior to Dec 1 2020 |  |  |  |  |  |
| - 0 | 8,990 (34.8%) | 7,390 (34.9%) | 7,716 (35.0%) | 8,563 (34.3%) | 0.01 |
| - 1 | 10,453 (40.4%) | 8,519 (40.2%) | 8,853 (40.1%) | 10,219 (40.9%) | 0.02 |
| - 2+ | 6,426 (24.8%) | 5,270 (24.9%) | 5,495 (24.9%) | 6,208 (24.8%) | 0.00 |
| Number of PCR tests taken from Dec 1 2020 to day of first vaccination |  |  |  |  |  |
| - 0 | 17,225 (66.6%) | 14,224 (67.2%) | 14,947 (67.7%) | 16,717 (66.9%) | 0.02 |
| - 1 | 6,447 (24.9%) | 5,207 (24.6%) | 5,328 (24.1%) | 6,195 (24.8%) | 0.01 |
| - 2+ | 2,197 (8.5%) | 1,748 (8.3%) | 1,789 (8.1%) | 2,078 (8.3%) | 0.01 |

For each outcome (i.e., SARS-CoV-2 infection and COVID-19 associated hospitalization, ICU admission, or death), we determined the baseline and breakthrough IRs for each cohort as described above. We calculated the incidence rate ratio (IRR) as the IR of a vaccinated cohort (mRNA-1273 or BNT162b2) divided by the IR of the unvaccinated cohort. The 95% confidence interval (CI) of the IRR was calculated using an exact method described previously.^12^ Effectiveness was then defined as 100 x (1 - IRR). The baseline (i.e., one week after first dose) IRR and effectiveness estimate for each outcome were included as controls to verify that the cohorts being compared were at similar risk for the given outcome at the time of study enrollment (i.e., the actual or hypothetical date of first vaccination).

We also performed Kaplan-Meier analysis to compare the cumulative incidence of SARS-CoV-2 infection or COVID-19 associated complications between vaccinated and unvaccinated individuals. Cumulative incidence at time *t* is the estimated proportion of individuals who experience the outcome on or before time *t* (i.e., 1 minus the standard Kaplan-Meier survival estimate). To analyze the effectiveness of full vaccination, we considered cumulative incidence from 14 days after the actual or hypothetical date of second vaccination. Statistical significance was assessed with the log rank test, and a p-value less than 0.05 was considered significant.^13^

### Assessing longitudinal effectiveness of mRNA-1273 and BNT162b2

We calculated monthly estimates of vaccine effectiveness against SARS-CoV-2 infection and COVID-19 associated hospitalization (as defined above) among matched unvaccinated and vaccinated individuals from Minnesota from February through July 2021. Effectiveness in each month was calculated as described above (i.e., by comparing the IRs of each vaccinated cohort to IRs of the unvaccinated cohort). For a given month, IRs for each group were calculated by dividing the number of individuals experiencing the outcome during that month by the total number of at-risk person-days contributed by fully vaccinated individuals in that month. Here, individuals contributed at-risk person days from the first day of the month or 14 days after their actual or hypothetical second vaccine dose (whichever came later) until (i) they experienced the outcome, (ii) they died, or (iii) the last day of the month (whichever came first).

### Comparing breakthrough infection incidence rates in the matched vaccinated cohorts

We determined the IRs of breakthrough infections in each matched vaccinated cohort (i.e., IR_mRNA-1273_ and IR_BNT162b2_) and computed the IRR as IR_mRNA-1273_ divided by IR_BNT162b2_. The 95% CI of the IRR was calculated using an exact method described previously.^12^ To verify that the matched cohorts were at similar risk for infection with SARS-CoV-2 at the time of study entry (i.e., date of first vaccine dose), we calculated the IRR of positive SARS-CoV-2 tests in the week after the first vaccine dose. An IRR was considered significantly different if its 95% CI did not include 1. This was performed for each state separately (Minnesota, Florida, Wisconsin, Arizona, Iowa) and for all states combined together. Note that unlike the effectiveness analyses described above (which was performed using exclusively individuals from Minnesota), we were able to perform this analysis across all contributing states because it does not require comparison against an unvaccinated cohort.

We also compared the cumulative incidence of breakthrough infections between the matched vaccinated cohorts from Minnesota using Kaplan-Meier analysis as described above.

### Comparing breakthrough infection-associated hospitalization, ICU admission, and mortality in the vaccinated matched cohorts

We defined breakthrough infection-associated hospitalization or ICU admission as hospitalization or ICU admission within 21 days of an individual’s first positive SARS-CoV-2 PCR test (i.e., COVID-19 diagnosis), where COVID-19 diagnosis occurred at least 14 days after the second vaccine dose. Breakthrough infection-associated death was defined similarly, except that we considered a 28 day window after the initial COVID-19 diagnosis (rather than 21 days).

We determined the IRs of each breakthrough infection-associated event (hospitalization, ICU admission, and death) in each matched vaccinated cohort and computed IRRs as was described above for the analysis of breakthrough infections themselves. We also calculated the IRR of each event in the one week after the first vaccine dose to verify that the cohorts were at similar risk for these events at the time of study entry. An IRR was considered significant if its 95% CI did not include 1.

To determine whether there were differences in the rates of disease severity outcomes between the mRNA-1273 and BNT162b2 cohorts *given* the diagnosis of a breakthrough infection, we also determined the 21-day hospitalization and ICU admission rates along with the 28-day mortality rate among patients who (i) experienced a breakthrough infection and (ii) contributed adequate follow-up time after their first positive SARS-CoV-2 PCR test for inclusion in the analysis (i.e., 21 or 28 days). These cumulative incidences were compared by calculating the risk ratio with a 95% CI and the Fisher exact test p-value. The rates were considered significantly different if the risk ratio 95% CI did not include 1 and the p-value was less than 0.05.

### Comparing potential complications experienced by patients with breakthrough infections

For all patients from the matched cohorts who experienced breakthrough infections (n = 106 for mRNA-1273; n = 220 for BNT162b2), we extracted potential complications from clinical notes of the EHR using an augmented curation BERT model trained to classify disease diagnosis.^14^ Specifically, this model classifies the sentiment of phenotype-containing sentences into one of three categories: Yes (i.e., positive diagnosis for disease X), No (i.e., ruled out diagnosis for disease X), or Maybe (e.g., family history or suspected diagnosis of disease X). This model was previously trained and validated to perform this type of classification task on a set of 18,490 sentences from clinical notes, and the model achieves an out-of-sample accuracy of 93.6% and precision/recall values over 95%.

For each patient, this model was applied to any clinical note in the 180 days prior to or 30 days after the first positive SARS-CoV-2 PCR test. The following phenotypes were considered as potential complications: acute respiratory distress syndrome/acute lung injury (ARDS/ALI), acute kidney injury, anemia, cardiac arrest, cardiac arrhythmias, disseminated intravascular coagulation, heart failure, hyperglycemia, hypertension, myocardial infarction, pleural effusion, pulmonary embolism, respiratory failure, sepsis, septic shock, stroke/cerebrovascular accident, venous thromboembolism, encephalopathy/delirium, and numbness. Importantly, while this model allows for the classification of diagnostic sentiment at the sentence level, it does not assess the time of phenotype onset and thereby does not indicate whether the given phenotype was caused by and/or is directly related to COVID-19. For example, if a sentence from an EHR note written 10 days after COVID-19 diagnosis suggests a positive diagnosis of hypertension, it is possible that this refers to pre-existing hypertension (e.g., a comorbidity which has continued through the current time rather than a complication) or to new-onset hypertension (e.g., a true potential COVID-19 complication).

After curating the clinical notes, we calculated IRs of each potential complication in the mRNA-1273 and BNT162b2 breakthrough infection cohorts during the COVID-19 associated interval (defined as days -3 to +30 relative to first positive PCR test). IRs were defined as the number of patients who experienced the complication divided by the total number of at-risk person-days contributed by the cohort. Patients contributed at-risk person days starting three days prior to their breakthrough diagnosis and extending until (i) they experienced the complication, (ii) they died, (iii) they reached day 30 after diagnosis, or (iv) the study period ended. If a patient had experienced the given complication in the pre-COVID interval (days -180 to -3 relative to first positive PCR test), then the phenotype was considered a likely pre-existing comorbidity or past medical event; therefore, such a patient was considered ineligible to experience the complication in the COVID-19 associated interval and would contribute no at-risk person days for that complication. We then calculated the IRR for each complication as the IR in the mRNA-1273 breakthrough cohort divided by the IR in the BNT162b2 breakthrough cohort. An IRR was considered significant if its 95% CI did not include 1.

### Assessing longitudinal prevalence of SARS-CoV-2 variants

Genomic sequence data from the GISAID initiative was used to estimate the longitudinal prevalence of SARS-CoV-2 variants in the states from which cohorts were derived (Minnesota, Arizona, Florida, Iowa, and Wisconsin).^15^ Specifically, we quantified the prevalence of the Pango lineages corresponding to CDC-labeled variants of interest (VOIs) and variants of concern (VOCs) in each state during approximately 15-day intervals (i.e., twice per month). For a given variant, prevalence was calculated as the number of sequences corresponding to that variant deposited in that state over the 15-day interval divided by the total number of sequences deposited in that state during the same interval, multiplied by 100. A total of 43,319 SARS-CoV-2 genome sequences collected between December 2020 and July 2021 were included in this analysis. The total number of deposited sequences split by state was as follows: Florida - 20,284; Minnesota - 15,485; Wisconsin - 3,853; Arizona - 3,263; Iowa - 434.

### IRB approval for human subjects research

This study was reviewed and approved by the Mayo Clinic Institutional Review Board (IRB 20-003278) as a minimal risk study. Subjects were excluded if they did not have a research authorization on file. The approved IRB was titled: Study of COVID-19 patient characteristics with augmented curation of Electronic Health Records (EHR) to inform strategic and operational decisions with the Mayo Clinic. The study was deemed exempt by the Mayo Clinic Institutional Review Board and waived from consent. The following resource provides further information on the Mayo Clinic Institutional Review Board and adherence to basic ethical principles underlying the conduct of research, and ensuring that the rights and well-being of potential research subjects are adequately protected: www.mayo.edu/research/institutional-review-board/overview.

## Results

From January to July 2021 in Minnesota, the effectiveness estimates of mRNA-1273 and BNT162b2 in preventing SARS-CoV-2 infection with onset at least 14 days after the second dose were 86% (95% CI: 81-90.6%, p=1.6×10^−42^) and 76% (95% CI: 69-81%, p=1.3×10^−31^), respectively (**Figure 1, Table 2, Figure S2A**). Full vaccination with either vaccine was also highly effective against COVID-19 associated hospitalization (mRNA-1273: 91.6%, 95% CI: 81-97%, p=8.3×10^−14^; BNT162b2: 85%, 95% CI: 73-93%, p=3.8×10^−12^), ICU admission (mRNA-1273: 93.3%, 95% CI: 57-99.8%, p=5.0×10^−4^; BNT162b2: 87%, 95% CI:46-98.6%, p=1.2×10^−3^), and death (no deaths in either cohort) (**Table 2, Figure S2B-C**).

**Table 2.** Vaccine effectiveness against SARS-CoV-2 infection and other COVID-19 associated outcomes in Minnesota. Incidence is calculated as the number of individuals experiencing the given outcome per 1000 at-risk person-days. The columns are: **(1) Time Period:** Time period relative to first or second vaccine dose; **(2)** Outcome: The defined COVID-19 related outcome, including a positive SARS-CoV-2 test or COVID-19 associated hospitalization, ICU admission, or death. **(3-5) Incidence Rates:** Number of individuals with in the cohort experiencing the outcome in the time period, divided by the number of at-risk person-days for the cohort in the time period; in brackets and parentheses, the number of cases per 1000 person-days and the number of individuals contributing at-risk person-days. **(6-8) Incidence Rate Ratio:** Incidence Rate of the given vaccinated cohort divided by the Incidence Rate of the unvaccinated cohort, along with the exact 95% confidence interval. Vaccine effectiveness is calculated as 100 x (1-IRR). (8) **Incidence Rate Ratio:** Incidence Rate of the mRNA-1273 cohort divided by the Incidence Rate of the BNT=162b2 cohort, along with the exact 95% confidence interval.^12^

| Time Period | Outcome | mRNA-1273<br>Incidence<br>Rate<br>Events/Person-Days<br>[Per 1000<br>Person-Days]<br>(# Individuals<br>Contributing) | BNT162b2<br>Incidence<br>Rate<br>Events/Person-Days<br>[Per 1000<br>Person-Days]<br>(# Individuals<br>Contributing) | Unvaccinated<br>Incidence<br>Rate<br>Events/Person-Days<br>[Per 1000<br>Person-Days]<br>(# Individuals<br>Contributing) | IRR<br>mRNA-1273<br>/ Unvax | IRR<br>BNT162b2 /<br>Unvax | IRR<br>mRNA-1273<br>/ BNT162b2 |
| --- | --- | --- | --- | --- | --- | --- | --- |
| Days 1-7<br>following first<br>dose | Positive<br>SARS-CoV-2<br>PCR Test | 74/180810<br>[0.41] (n =<br>25869) | 58/180675<br>[0.32] (n =<br>25869) | 69/180614<br>[0.38] (n =<br>25869) | 1.1 (0.76, 1.5) | 0.84 (0.58,<br>1.2) | 1.3 (0.89, 1.8) |
|  | COVID-19<br>Associated<br>Hospitalization | 6/180937<br>[0.033] (n =<br>25869) | 2/180812<br>[0.011] (n =<br>25869) | 8/180798<br>[0.044] (n =<br>25869) | 0.75 (0.21, 2.5) | 0.25 (0.026,<br>1.3) | 3 (0.54, 30) |
|  | COVID-19<br>Associated<br>ICU<br>Admission | 0/180951 [0] (n<br>= 25869) | 0/180814 [0] (n<br>= 25869) | 2/180812<br>[0.011] (n =<br>25869) | 0 (0, 5.3) | 0 (0, 5.3) | N/A |
|  | COVID-19<br>Associated<br>Death | 0/180951 [0] (n<br>= 25869) | 0/180814 [0] (n<br>= 25869) | 0/180819 [0] (n<br>= 25869) | N/A | N/A | N/A |
| On or after<br>14 days<br>following the<br>second dose | Positive<br>SARS-CoV-2<br>PCR Test | 38/2214873.0<br>[0.017] (n =<br>21179) | 72/2332005.0<br>[0.031] (n =<br>22064) | 321/2526895.0<br>[0.13] (n =<br>24990) | 0.14 (0.094,<br>0.19) | 0.24 (0.19,<br>0.31) | 0.56 (0.36,<br>0.83) |
|  | COVID-19<br>Associated<br>Hospitalization | 6/2215483.0<br>[0.0027] (n =<br>21187) | 11/2333145.0<br>[0.0047] (n =<br>22085) | 82/2532948.0<br>[0.032] (n =<br>25083) | 0.084 (0.03,<br>0.19) | 0.15 (0.07,<br>0.27) | 0.57 (0.17, 1.7) |
|  | COVID-19<br>Associated<br>ICU<br>Admission | 1/2215536.0<br>[0.00045] (n =<br>21187) | 2/2333352.0<br>[0.00086] (n =<br>22090) | 17/2534192.0<br>[0.0067] (n =<br>25097) | 0.067 (0.0016,<br>0.43) | 0.13 (0.014,<br>0.54) | 0.53 (0.0089,<br>10) |
|  | COVID-19<br>Associated<br>Death | 0/2215773.0 [0]<br>(n = 21187) | 0/2333860.0 [0]<br>(n = 22092) | 4/2537030.0<br>[0.0016] (n =<br>25101) | 0 (0, 1.7) | 0 (0, 1.6) | N/A |

**Figure 1.**
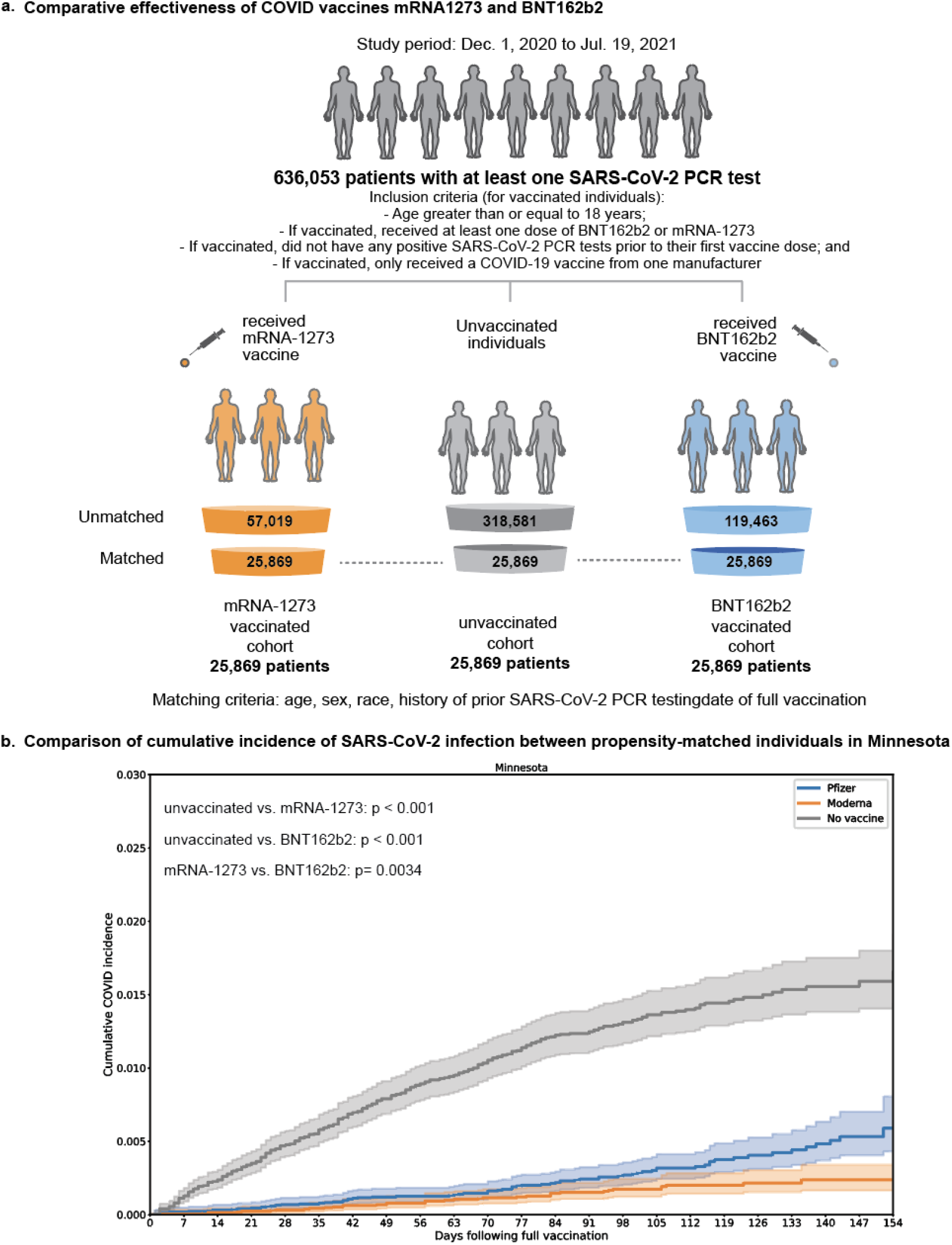
Study Overview. (A) Derivation of matched vaccinated and unvaccinated cohorts to compare the effectiveness of the mRNA COVID-19 vaccines mRNA1273 and BNT162b2. The matching process yielded 25,689 triples of individuals (one unvaccinated, one vaccinated with mRNA-1273, one vaccinated with BNT162b2) from Minnesota who were matched on the basis of age, sex, race, ethnicity, history of prior SARS-CoV-2 PCR testing, and date of vaccination. (B) With the cohorts described in (A), we assessed the overall effectiveness of each vaccine by comparing the cumulative incidence of infection in each vaccinated cohort compared to the matched unvaccinated cohort. We also assessed the relative effectiveness of each vaccine (i.e., incidence rate of infection in the mRNA-1273 cohort compared to the BNT162b2 cohort).

These estimates of effectiveness against infection (86% and 76%) were lower than those that we previously observed in the Mayo Clinic Health System through April 20, 2021 (mRNA-1273: 93.3%, 95% CI: 85.7-97.4%; BNT162b2: 86.1%, 95% CI: 82.4-89.1%).^6^ We thus analyzed the effectiveness of full vaccination longitudinally on a monthly basis starting in February 2021 (see **Methods**). In the context of increasing cases in Minnesota during July (**Figure S3**), the effectiveness against infection was lower for mRNA-1273 (76%, 95% CI: 58-87%) compared to prior months, with an even more pronounced reduction for BNT162b2 (42%, 95% CI: 13-62%) (**Figure 2A**; **Table 3**). Importantly, the effectiveness of mRNA-1273 and BNT162b2 against COVID-19 associated hospitalization has remained more consistently high (**Figure 2B, Table 4**). Of note, July corresponds to the time during which the Delta variant has risen to prominence in Minnesota (**Figure 2C**).

**Table 3.** Longitudinal analysis of vaccine effectiveness against breakthrough infections in Minnesota, split by month.

| **Month** | **mRNA-1273**<br>**Incidence Rate**<br>**Events/Person-Days**<br>**[Per 1000 Person-Days]**<br>**(# Individuals**<br>**Contributing)** | **BNT162b2**<br>**Incidence Rate**<br>**Events/Person-Days**<br>**[Per 1000 Person-Days]**<br>**(# Individuals**<br>**Contributing)** | **Unvaccinated**<br>**Incidence Rate**<br>**Events/Person-Days**<br>**[Per 1000**<br>**Person-Days]**<br>**(# Individuals**<br>**Contributing)** | **IRR**<br>**mRNA-1273 /**<br>**Unvax** | **IRR**<br>**BNT162b2**<br>**/ Unvax** | **IRR**<br>**mRNA-1273 /**<br>**BNT162b2** |
| --- | --- | --- | --- | --- | --- | --- |
| February | 0/14106.0 [0] (n = 1574) | 0/15953.0 [0] (n = 1552) | 1/15813.0 [0.063] (n = 1595) | 0 (0, 44) | 0 (0, 39) | N/A |
| March | 3/123184.0 [0.024] (n = 6956) | 4/135794.0 [0.029] (n = 6590) | 37/137738.0 [0.27] (n = 7411) | 0.091 (0.018, 0.29) | 0.11 (0.028, 0.31) | 0.83 (0.12, 4.9) |
| April | 7/302898.0 [0.023] (n = 14463) | 11/330239.0 [0.033] (n = 15129) | 93/343015.0 [0.27] (n = 16321) | 0.085 (0.033, 0.18) | 0.12 (0.059, 0.23) | 0.69 (0.23, 2) |
| May | 5/508598.0 [0.0098] (n = 18449) | 13/533743.0 [0.024] (n = 19503) | 82/575635.0 [0.14] (n = 21212) | 0.069 (0.022, 0.17) | 0.17 (0.087, 0.31) | 0.4 (0.11, 1.2) |
| June | 8/575955.0 [0.014] (n = 20583) | 4/598933.0 [0.0067] (n = 21411) | 24/659357.0 [0.036] (n = 23780) | 0.38 (0.15, 0.88) | 0.18 (0.046, 0.53) | 2.1 (0.56, 9.4) |
| July | 15/627756.0 [0.024] (n = 21079) | 38/652459.0 [0.058] (n = 21946) | 73/724645.0 [0.1] (n = 24444) | 0.24 (0.13, 0.42) | 0.58 (0.38, 0.87) | 0.41 (0.21, 0.76) |

**Table 4.** Longitudinal analysis of vaccine effectiveness against hospitalizations associated with breakthrough infections in Minnesota, split by month.

| **Month** | **mRNA-1273**<br>**Incidence Rate**<br>**Events/Person-Days**<br>**[Per 1000 Person-Days]**<br>**(# Individuals**<br>**Contributing)** | **BNT162b2**<br>**Incidence Rate**<br>**Events/Person-Days**<br>**[Per 1000 Person-Days]**<br>**(# Individuals**<br>**Contributing)** | **Unvaccinated**<br>**Incidence**<br>**Rate**<br>**Events/Person**<br>**-Days**<br>**[Per 1000**<br>**Person-Days]**<br>**(# Individuals**<br>**Contributing)** | **IRR**<br>**mRNA-1273**<br>**/ Unvax** | **IRR**<br>**BNT162b2 /**<br>**Unvax** | **IRR**<br>**mRNA-1273 /**<br>**BNT162b2** |
| --- | --- | --- | --- | --- | --- | --- |
| February | 0/14117.0 [0] (n = 1575) | 0/15974.0 [0] (n = 1554) | 0/15848.0 [0] (n = 1601) | N/A | N/A | N/A |
| March | 1/123233.0 [0.0081] (n = 6958) | 1/135930.0 [0.0074] (n = 6594) | 9/138478.0 [0.065] (n = 7435) | 0.12 (0.0028, 0.9) | 0.11 (0.0026, 0.82) | 1.1 (0.014, 87) |
| April | 1/303102.0 [0.0033] (n = 14469) | 2/330648.0 [0.006] (n = 15144) | 20/346174.0 [0.058] (n = 16422) | 0.057 (0.0014, 0.36) | 0.1 (0.012, 0.43) | 0.55 (0.0092, 10) |
| May | 1/509149.0 [0.002] (n = 18465) | 3/534837.0 [0.0056] (n = 19536) | 25/582284.0 [0.043] (n = 21411) | 0.046 (0.0011, 0.28) | 0.13 (0.025, 0.43) | 0.35 (0.0067, 4.4) |
| June | 0/576677.0 [0] (n = 20603) | 1/600226.0 [0.0017] (n = 21455) | 7/667073.0 [0.01] (n = 24039) | 0 (0, 0.8) | 0.16 (0.0035, 1.2) | 0 (0, 41) |
| July | 3/628674.0 [0.0048] (n = 21107) | 4/654236.0 [0.0061] (n = 21994) | 18/733732.0 [0.025] (n = 24721) | 0.19 (0.037, 0.67) | 0.25 (0.061, 0.76) | 0.78 (0.11, 4.6) |

**Figure 2.**
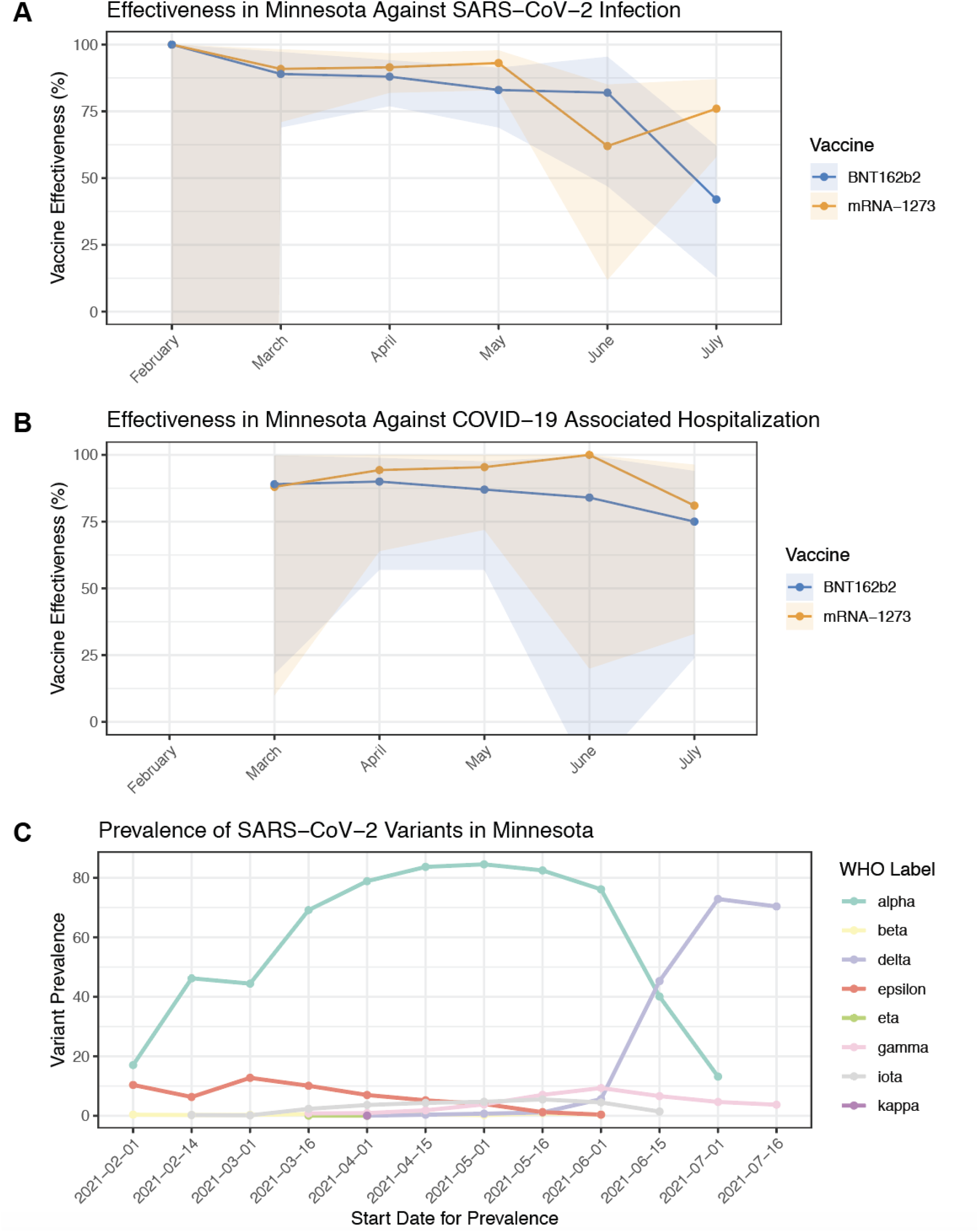
Longitudinal analysis of vaccine effectiveness and SARS-CoV-2 variant landscape in Minnesota. (A) Monthly estimates of vaccine effectiveness against SARS-CoV-2 infection for mRNA-1273 and BNT162b2 in Minnesota, calculated by comparing the incidence rates of positive testing in each vaccinated cohort during that month (i.e., not cumulative) to the incidence rate of positive testing in the matched unvaccinated cohort during that month. (B) Monthly estimates of vaccine effectiveness against COVID-19 associated hospitalization for mRNA-1273 and BNT162b2 in Minnesota, calculated as described in (A) but considering hospitalization within 21 days of infection as the outcome rather than infection alone. (C) Prevalence of SARS-CoV-2 variants in Minnesota, assessed twice monthly during the study period. In (A) and (B), points correspond to point estimates for monthly vaccine effectiveness, and shaded regions represent the corresponding 95% CIs.

In addition to the changing effectiveness against infection over time, we noted that the 95% confidence intervals of the estimates for effectiveness against infection did not overlap (mRNA-1273: 81-90.6%; BNT162b2: 69-81%) (**Table 2**). The incidence rate of breakthrough infections over the study duration was significantly lower in the mRNA-1273 cohort (IR_mRNA-1273_: 0.017, IR_BNT162b2_: 0.031; IRR = 0.56, 95% CI: 0.36-0.83) despite similar baseline infection risks in the week after the first vaccine dose (IRR = 1.3; 95% CI: 0.89-1.8) (**Table 2**). Kaplan-Meier analysis indicates a difference in cumulative breakthrough infection incidence between the vaccinated cohorts (p=3.4×10^−3^; **Figure S2A**). On the other hand, the mRNA-1273 and BNT162b2 cohorts had similar rates of hospitalization (IRR: 0.57, 95% CI: 0.17-1.7, p=0.30), ICU admission (IRR: 0.53, 95% CI: 0.0089-10, p=0.59), and death (no events in either cohort) (**Table 2** and **Figures S2B-C**).

To validate these findings from Minnesota regarding the comparative effectiveness of mRNA-1273 and BNT162b2, we next compared the rates of breakthrough infections between matched individuals vaccinated with mRNA-1273 versus BNT162b2 in other states from the Mayo Clinic Health System (Wisconsin, Arizona, Florida, and Iowa). In most states, individuals vaccinated with mRNA-1273 were less likely to experience breakthrough infections over the duration of the study period (IRR_Florida_: 0.42, 95% CI: 0.28-0.62; IRR_Wisconsin_: 0.52, 95% CI: 0.26-1.0; IRR_Arizona_: 0.39, 95% CI: 0.15-0.92; IRR_Iowa_: 0.0, 95% CI: 0.0-1.6) (**Table 5**). Considering all states together, mRNA-1273 conferred a two-fold risk reduction against breakthrough infection compared to BNT162b2 (IRR = 0.50, 95% CI: 0.39-0.64) (**Table 5**). A monthly comparative analysis highlighted that the difference in infection risk was strongest in July (IRR: 0.44; 95% CI: 0.32-0.60) (**Table 6**), during which the Delta variant has risen to over 50% prevalence in each represented state (**Figure S4**). This was especially prominent during the recent case surge in Florida (**Figure S3**), where the risk of infection in July after full vaccination with mRNA-1273 was about 60% lower than after full vaccination with BNT162b2 (IRR: 0.39, 95% CI: 0.24-0.62) (**Table 5**). Across all states, individuals vaccinated with mRNA-1273 also experienced COVID-19 associated hospitalizations at approximately half the rate of individuals vaccinated with BNT162b2 (IRR: 0.51, 95% CI: 0.29-0.88), while there was no significant difference between the cohorts regarding the incidence rates of COVID-19 associated ICU admission (IRR: 0.75, 95% CI: 0.19-2.7) or mortality (**Table 7**).

**Table 5.**
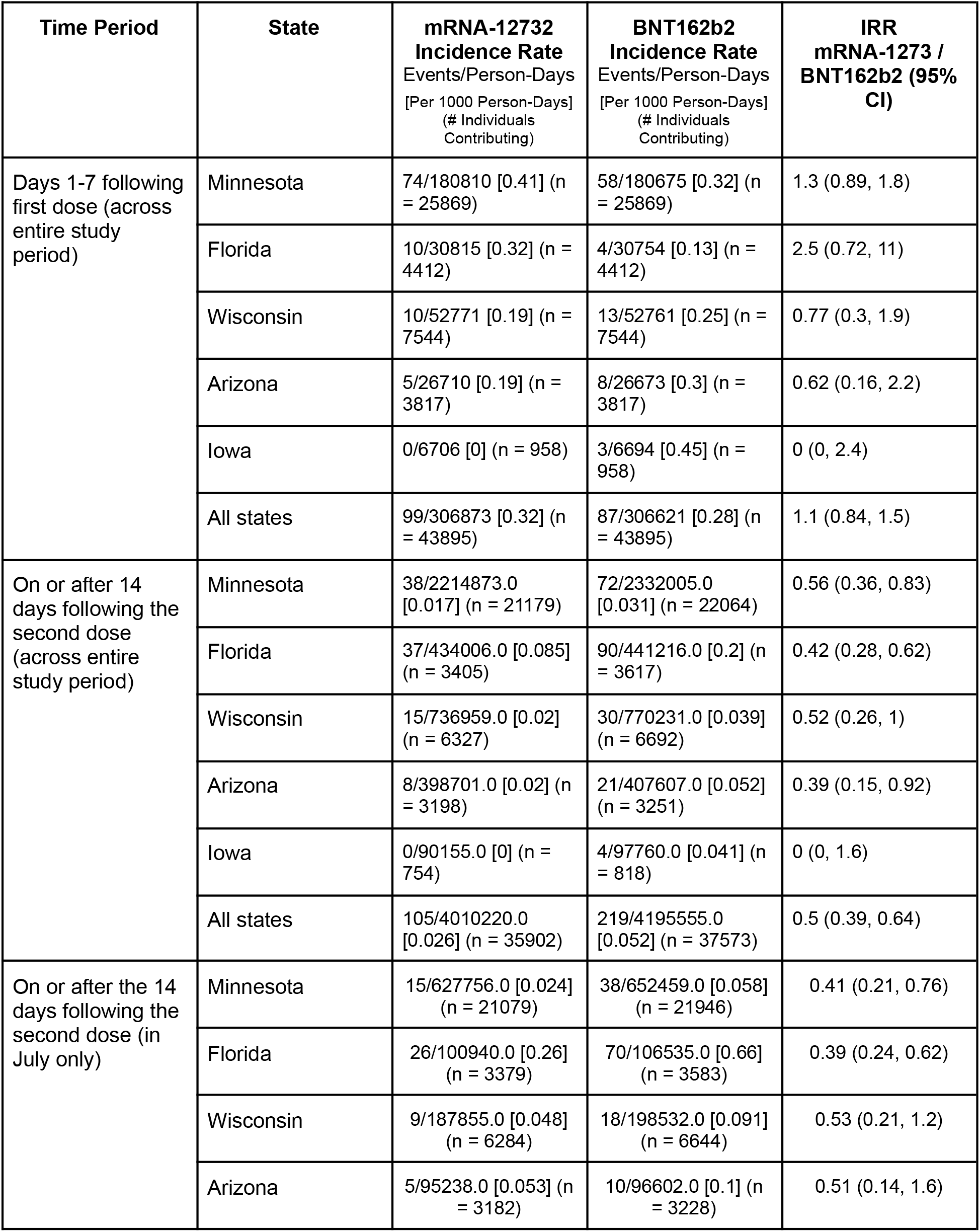
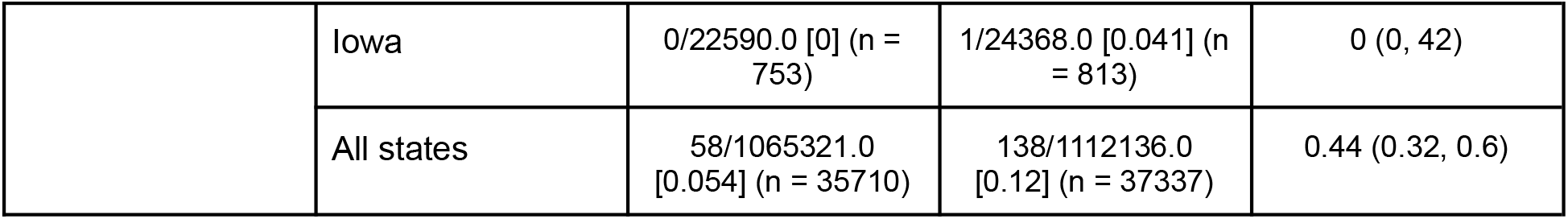
Comparison of incidence rates of positive SARS-CoV-2 testing between mRNA-1273 cohort versus BNT162b2 cohort within each individual state and across all states.

| **Time Period** | **State** | **mRNA-12732**<br>**Incidence Rate**<br>Events/Person-Days<br>[Per 1000 Person-Days]<br>(# Individuals<br>Contributing) | **BNT162b2**<br>**Incidence Rate**<br>Events/Person-Days<br>[Per 1000 Person-Days]<br>(# Individuals<br>Contributing) | **IRR**<br>**mRNA-1273 /**<br>**BNT162b2 (95%**<br>**CI)** |
| --- | --- | --- | --- | --- |
| Days 1-7 following first dose (across entire study period) | Minnesota | 74/180810 [0.41] (n = 25869) | 58/180675 [0.32] (n = 25869) | 1.3 (0.89, 1.8) |
|  | Florida | 10/30815 [0.32] (n = 4412) | 4/30754 [0.13] (n = 4412) | 2.5 (0.72, 11) |
|  | Wisconsin | 10/52771 [0.19] (n = 7544) | 13/52761 [0.25] (n = 7544) | 0.77 (0.3, 1.9) |
|  | Arizona | 5/26710 [0.19] (n = 3817) | 8/26673 [0.3] (n = 3817) | 0.62 (0.16, 2.2) |
|  | Iowa | 0/6706 [0] (n = 958) | 3/6694 [0.45] (n = 958) | 0 (0, 2.4) |
|  | All states | 99/306873 [0.32] (n = 43895) | 87/306621 [0.28] (n = 43895) | 1.1 (0.84, 1.5) |
| On or after 14 days following the second dose (across entire study period) | Minnesota | 38/2214873.0 [0.017] (n = 21179) | 72/2332005.0 [0.031] (n = 22064) | 0.56 (0.36, 0.83) |
|  | Florida | 37/434006.0 [0.085] (n = 3405) | 90/441216.0 [0.2] (n = 3617) | 0.42 (0.28, 0.62) |
|  | Wisconsin | 15/736959.0 [0.02] (n = 6327) | 30/770231.0 [0.039] (n = 6692) | 0.52 (0.26, 1) |
|  | Arizona | 8/398701.0 [0.02] (n = 3198) | 21/407607.0 [0.052] (n = 3251) | 0.39 (0.15, 0.92) |
|  | Iowa | 0/90155.0 [0] (n = 754) | 4/97760.0 [0.041] (n = 818) | 0 (0, 1.6) |
|  | All states | 105/4010220.0 [0.026] (n = 35902) | 219/4195555.0 [0.052] (n = 37573) | 0.5 (0.39, 0.64) |
| On or after the 14 days following the second dose (in July only) | Minnesota | 15/627756.0 [0.024] (n = 21079) | 38/652459.0 [0.058] (n = 21946) | 0.41 (0.21, 0.76) |
|  | Florida | 26/100940.0 [0.26] (n = 3379) | 70/106535.0 [0.66] (n = 3583) | 0.39 (0.24, 0.62) |
|  | Wisconsin | 9/187855.0 [0.048] (n = 6284) | 18/198532.0 [0.091] (n = 6644) | 0.53 (0.21, 1.2) |
|  | Arizona | 5/95238.0 [0.053] (n = 3182) | 10/96602.0 [0.1] (n = 3228) | 0.51 (0.14, 1.6) |
|  | Iowa | 0/22590.0 [0] (n = 753) | 1/24368.0 [0.041] (n = 813) | 0 (0, 42) |
|  | All states | 58/1065321.0 [0.054] (n = 35710) | 138/1112136.0 [0.12] (n = 37337) | 0.44 (0.32, 0.6) |

**Table 6.** Incidence rates of breakthrough infections across entire matched cohorts (all states included), split by month.

| **Month** | **mRNA-1273 Incidence Rate**<br>Events/Person-Days<br>[Per 1000 Person-Days]<br>(# Individuals Contributing) | **BNT162b2 Incidence Rate**<br>Events/Person-Days<br>[Per 1000 Person-Days]<br>(# Individuals Contributing) | **IRR**<br>**mRNA-1273 / BNT162b2** |
| --- | --- | --- | --- |
| February | 0/30026.0 [0] (n = 3770) | 0/31074.0 [0] (n = 3351) | N/A |
| March | 6/268901.0 [0.022] (n = 15373) | 8/281110.0 [0.028] (n = 14188) | 0.78 (0.22, 2.6) |
| April | 12/612527.0 [0.02] (n = 26911) | 24/647037.0 [0.037] (n = 28001) | 0.53 (0.24, 1.1) |
| May | 14/921359.0 [0.015] (n = 32862) | 18/965133.0 [0.019] (n = 34577) | 0.81 (0.38, 1.7) |
| June | 15/997221.0 [0.015] (n = 35193) | 27/1040827.0 [0.026] (n = 36789) | 0.58 (0.29, 1.1) |
| July | 58/1065321.0 [0.054] (n = 35710) | 138/1112136.0 [0.12] (n = 37337) | 0.44 (0.32, 0.6) |

**Table 7.** Incidence rates of COVID-19 associated complications across entire matched cohorts (all states included).

| **Time Period** | **Outcome** | **mRNA-1273 Incidence Rate**<br>Events/Person-Days<br>[Per 1000 Person-Days]<br>(# Individuals Contributing) | **BNT162b2 Incidence Rate**<br>Events/Person-Days<br>[Per 1000 Person-Days]<br>(# Individuals Contributing) | **IRR mRNA-1273 / BNT162b2** |
| --- | --- | --- | --- | --- |
| Days 1-7 following first dose | COVID-19 Associated Hospitalization | 8/307037 [0.026] (n = 43895) | 8/306843 [0.026] (n = 43895) | 1 (0.33, 3.1) |
|  | COVID-19 Associated ICU Admission | 1/307053 [0.0033] (n = 43895) | 0/306850 [0] (n = 43895) | inf (0.026, inf) |
|  | COVID-19 Associated Death | 0/307054 [0] (n = 43895) | 0/306850 [0] (n = 43895) | N/A |
| On or after 14 days following the second dose | COVID-19 Associated Hospitalization | 21/4011550.0 [0.0052] (n = 35914) | 43/4198278.0 [0.01] (n = 37603) | 0.51 (0.29, 0.88) |
|  | COVID-19 Associated ICU Admission | 5/4011773.0 [0.0012] (n = 35915) | 7/4198947.0 [0.0017] (n = 37610) | 0.75 (0.19, 2.7) |
|  | COVID-19 Associated Death | 1/4012246.0 [0.00025] (n = 35915) | 0/4199985.0 [0] (n = 37612) | Inf (0.027, Inf) |

Finally, we examined whether there were differences in the conditional risk of experiencing complications or severe disease given the diagnosis of a breakthrough infection. Augmented curation of clinical notes (see **Methods**) showed that all assayed complications were experienced at similar rates between mRNA-1273 and BNT162b2 breakthrough patients (**Table 8**). There were also no significant differences in the rates of 21-day hospitalization (mRNA-1273: 11/48 [22.9%]; BNT162b2: 27/103 [26.2%]; Risk Ratio = 0.87, 95% CI: 0.49-1.6; p = 0.84), 21-day ICU admission (mRNA-1273: 2/48 [4.2%]; BNT162b2: 5/103 [4.9%]; Risk Ratio = 0.86, 95% CI: 0.22-4.1; p = 1.0), or 28-day mortality (mRNA-1273: 1/48 [2.1%]; BNT162b2: 0/87 [0.0%]; Risk Ratio = Infinity, 95% CI: 0.22-Infinity; p = 0.36) (**Table 9**).

**Table 8.** Incidence rates of potential COVID-19 associated complications in breakthrough patients. The columns are: **(1) Complications:** phenotypes that are written with positive-sentiment in the clinical notes and occur -3 to +30 days relative to COVID diagnosis and do not occur -180 to -4 days relative to COVID diagnosis; **(2) Incidence Rate of Complications in mRNA-1273 Breakthrough cases:** the number of mRNA-1273-vaccinated individuals experiencing the complication divided by the number of at-risk patient days contributed by mRNA-1273-vaccinated breakthrough cases; **(3) Incidence Rate of Complications in BNT162b2 Breakthrough cases:** the number of BNT162b2-vaccinated individuals experiencing the complication divided by the number of at-risk patient days contributed by BNT162b2-vaccinated breakthrough cases; **(4) IRR mRNA-1273 / BNT162b2:** the IR of the complication in the mRNA-1273 breakthrough cohort divided by the IR of the complication in the BNT162b2 breakthrough cohort.

| **Complication** | **Incidence Rate**<br>**mRNA-1273**<br>**(n = 106)**<br>**Events/Person-Days**<br>**[Per 1000 Person-Days]** | **Incidence Rate**<br>**BNT162b2**<br>**(n = 220)**<br>**Events/Person-Days**<br>**[Per 1000 Person-Days]** | **IRR mRNA-1273 /**<br>**BNT162b2**<br>**(exact 95% CI)** |
| --- | --- | --- | --- |
| ARD ALI | 4 / 2,206 [0.18%] | 10 / 4,335 [0.23%] | 0.79 (0.18, 2.73) |
| Acute kidney injury | 5 / 2,052 [0.24%] | 15 / 4,062 [0.37%] | 0.66 (0.19, 1.91) |
| Anemia | 9 / 1,729 [0.52%] | 8 / 3,853 [0.21%] | 2.51 (0.86, 7.47) |
| Cardiac arrest | 1 / 2,230 [0.045%] | 2 / 4,521 [0.044%] | 1.01 (0.02, 19.47) |
| Cardiac arrhythmias | 9 / 1,686 [0.53%] | 20 / 3,216 [0.62%] | 0.86 (0.34, 1.97) |
| Chronic fatigue syndrome | 1 / 2,233 [0.045%] | 0 / 4,591 [0%] | inf (0.05, inf) |
| Disseminated intravascular<br>coagulation | 0 / 2,238 [0%] | 0 / 4,591 [0%] | N/A |
| Encephalopathy Delirium | 2 / 2,148 [0.093%] | 4 / 4,360 [0.092%] | 1.01 (0.09, 7.08) |
| Heart failure | 5 / 2,043 [0.24%] | 7 / 4,149 [0.17%] | 1.45 (0.36, 5.31) |
| Hyperglycemia | 3 / 2,045 [0.15%] | 5 / 3,988 [0.13%] | 1.17 (0.18, 6.01) |
| Hypertension | 8 / 1,295 [0.62%] | 24 / 2,137 [1.1%] | 0.55 (0.21, 1.27) |
| Myocardial infarction | 2 / 2,143 [0.093%] | 5 / 4,364 [0.11%] | 0.81 (0.08, 4.98) |
| Numbness | 4 / 1,873 [0.21%] | 5 / 3,930 [0.13%] | 1.68 (0.33, 7.8) |
| Pleural effusion | 5 / 2,140 [0.23%] | 8 / 4,240 [0.19%] | 1.24 (0.32, 4.29) |
| Pulmonary embolism | 2 / 2,208 [0.091%] | 5 / 4,393 [0.11%] | 0.8 (0.08, 4.86) |
| Respiratory failure | 5 / 2,199 [0.23%] | 10 / 4,335 [0.23%] | 0.99 (0.26, 3.17) |
| Sepsis | 3 / 2,142 [0.14%] | 2 / 4,418 [0.045%] | 3.09 (0.35, 37.04) |
| Septic shock | 1 / 2,217 [0.045%] | 0 / 4,557 [0%] | inf (0.05, inf) |
| Stroke | 0 / 2,204 [0%] | 5 / 4,307 [0.12%] | 0.0 (0, 2.13) |
| Venous thromboembolism | 3 / 2,173 [0.14%] | 3 / 4,382 [0.068%] | 2.02 (0.27, 15.06) |

**Table 9.** 21-day hospitalization, 21-day ICU admission, and 28-day mortality rates among breakthrough cases from the matched mRNA-1273 and BNT162b2 cohorts. The columns are: **(1) Outcome:** the clinical metric of COVID-19 severity assessed in the given row; **(1) mRNA-1273 Breakthrough Cohort:** cumulative incidence of the given outcome among mRNA-1273 breakthrough cases; **(2) Prevalence in BNT162b2 Breakthrough Cohort:** cumulative incidence of the given outcome among mRNA-1273 breakthrough cases; **(3) Risk Ratio (95% CI):** for the given outcome, cumulative incidence in the mRNA-1273-vaccinated cohort divided by cumulative incidence in the BNT162b2-vaccinated cohort, along with the 95% confidence interval; **(4) Risk Ratio (95% CI):** for the given outcome, cumulative incidence in the mRNA-1273-vaccinated cohort divided by cumulative incidence in the BNT162b2-vaccinated cohort, along with the 95% confidence interval. **(5) Fisher Exact P-Value:** for the given outcome, the p-value from a Fisher exact test performed on a two-by-two table of vaccine group (mRNA-1273 versus BNT162b2) by outcome status (Yes versus No).

| **Outcome** | **mRNA-1273 Breakthrough Cohort** | **BNT162b2 Breakthrough Cohort** | **Risk Ratio (95% CI)** | **Fisher Exact P-Value** |
| --- | --- | --- | --- | --- |
| 21-Day Hospitalization | 11/48 (22.9%) | 27/103 (26.2%) | 0.87 (0.49, 1.6) | 0.84 |
| 21-Day ICU Admission | 2/48 (4.2%) | 5/103 (4.9%) | 0.86 (0.22, 4.1) | 1 |
| 28-Day Mortality | 1/48 (2.1%) | 0/87 (0.0%) | inf (0.22, inf) | 0.36 |

## Discussion

The occurrence of breakthrough infections and reports of diminished neutralization of emergent variants by vaccine-elicited sera mandate the continual monitoring of the comparative effectiveness and durability of COVID-19 vaccines.^8,9^ Overall, we find that in our study population from Minnesota, both vaccines strongly reduce the risk of SARS-CoV-2 infection and severe COVID-19, but individuals vaccinated with mRNA-1273 were about half as likely to experience breakthrough infections as individuals vaccinated with BNT162b2. This relative risk reduction conferred by mRNA-1273 was also observed in other states, including in Florida during a recent COVID-19 outbreak. The effectiveness of both vaccines, particularly BNT162b2, was lower in July compared to prior months. Finally, the rates of complications experienced by patients with breakthrough infections were similar between those vaccinated with mRNA-1273 or BNT162b2.

mRNA-1273 and BNT162b2 were originally designed, tested, and proven to reduce the burden of symptomatic disease, hospitalization, and death related to SARS-CoV-2 infection. This study further supports the effectiveness of both vaccines in doing so, even despite the evolution of more transmissible viral variants. It is important to realize that most vaccines are not 100% effective, particularly against asymptomatic infections. For example, the estimated effectiveness of seasonal influenza vaccines has ranged from 19-60% over the past decade.^16^ While COVID-19 mRNA vaccines have been shown to be drastically more effective than this, the occurrence of breakthrough infections is indeed still expected. We observed a pronounced reduction in the effectiveness of BNT162b2 coinciding with the surging prevalence of the Delta variant in the United States, but this temporal association does not imply causality, and there are likely several factors contributing to changes in vaccine effectiveness over time. Consistent with our findings, a previous test-negative case-control study found that full vaccination with BNT162b2 was less effective in preventing symptomatic infection with the Delta variant (88.0%, 95% CI: 85-90.1%) than with the Alpha variant (93.7%, 95% CI: 91.6-95.3%), although it was highly effective against both.^17^

Several factors could contribute to the observed differences in effectiveness of mRNA-1273 and BNT162b2. Although both are nucleoside-modified mRNA vaccines encoding the prefusion stabilized SARS-CoV-2 Spike protein, there are differences in the vaccination regimen and formulation.^18,19^ BNT162b2 is administered as 30μg/0.3mL (100 μg/mL) doses 21 days apart^20^ and the Moderna vaccine is administered as 100μg/0.5mL (200 μg/mL) doses 28 days apart.^21^ Assuming similar sized constructs, this means that each mRNA-1273 dose provides three times more mRNA copies of the Spike protein than BNT162b2, which could result in more effective priming of the immune response. There has not been a head-to-head comparison of the neutralizing antibody titers elicited by BNT162b2 versus mRNA-1273, but such a study could provide important context for our results. Certain adverse effects, such as myalgia and arthralgia, were observed more frequently after vaccination with mRNA-1273 than BNT162b2 in their respective clinical trials, and it can be speculated that this increased reactogenicity is paralleled by increased immunogenicity.^3,4^ Furthermore, there are differences in the lipid composition of the nanoparticles used for packaging the mRNA content of mRNA-1273 and BNT162b2. BNT162b2 has a lipid nanoparticle composed of ALC-0315, ALC-0159, distearolyphosphatidycholine (DSPC), and cholesterol whereas the lipid nanoparticle of mRNA-1273 is composed of SM-102, PEG-DMG, DSPC, and cholesterol.^22^ The structures of the cationic lipids (ALC-0315 and SM-102) in each formulation are shown in **Figure S5**.

There are some limitations of this study. First, these cohorts are not demographically representative of the American population (**Table 1, Table S1**), which may limit the generalizability of our findings. Similar real world clinical studies on larger and more diverse populations from various health systems are needed to more robustly compare the effectiveness of mRNA-1273 and BNT162b2. Second, although this study accounts for geographic variability by matching individuals from the same state, these conclusions should continue to be tested longitudinally throughout the United States and globally. Third, it is possible that our vaccine effectiveness estimates are impacted by unknown exposure risk variables which were missed in the matching procedure, although the similar risks for infection, hospitalization, ICU admission, and death in the week following the first dose suggest that all of the compared cohorts had similar baseline risks for the defined outcomes at the time of study enrollment. Finally, while we did observe a recent reduction in vaccine effectiveness in July, we did not analyze the risk of infection relative to the date of vaccination. The reduced effectiveness could be due to waning immunity over time, the dynamic landscape of SARS-CoV-2 variants, or other factors that were not considered here.

Our observational study suggests that while both mRNA COVID-19 vaccines strongly protect against infection and severe disease, there are differences in their real-world effectiveness relative to each other and relative to prior months of the pandemic. Larger studies with more diverse populations are warranted to guide critical pending public and global health decisions, such as the optimal timing for booster doses and which vaccines should be administered to individuals who have not yet received one dose. As we continue to vigilantly monitor longitudinal and comparative vaccine effectiveness in the coming months, this study emphasizes the importance of vaccination to reduce the risk of SARS-CoV-2 infection and its associated complications.

## Data Availability

After publication, the data will be made available upon reasonable requests to the corresponding author. A proposal with a detailed description of study objectives and the statistical analysis plan will be needed for evaluation of the reasonability of requests. Deidentified data will be provided after approval from the corresponding author and the Mayo Clinic.

## Acknowledgements

The authors thank Murali Aravamudan for his feedback on this manuscript.

## Supplementary Material

**Figure S1.**
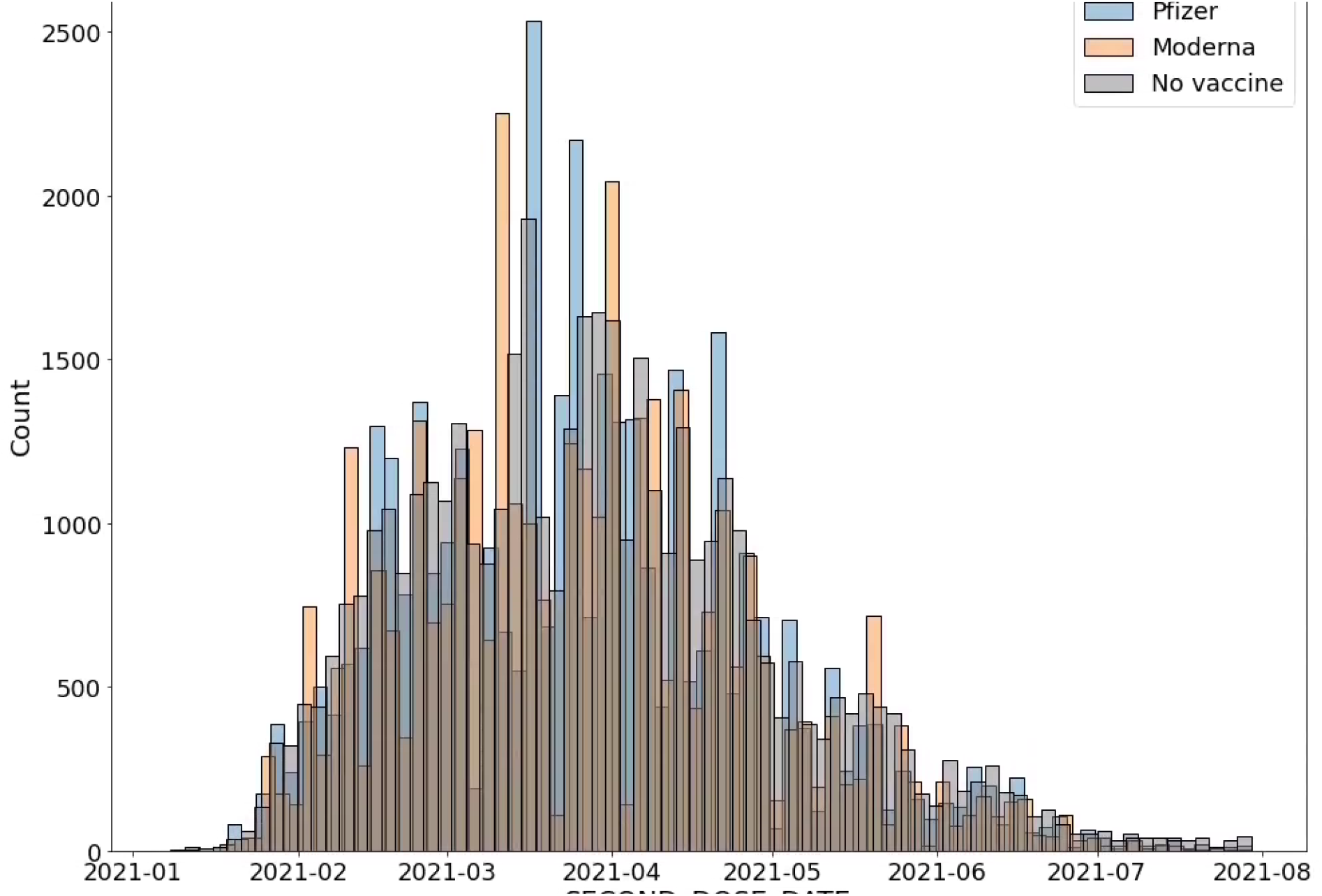
Date of actual or hypothetical second vaccine dose for the matched mRNA-1273, BNT162b2, and unvaccinated cohorts from Minnesota. The median date of second dose administration in the matched cohorts is March 31, 2021 for Pfizer, Apr 1, 2021 for Moderna, and April 2, 2021 for the matched unvaccinated cohort.

**Figure S2.**
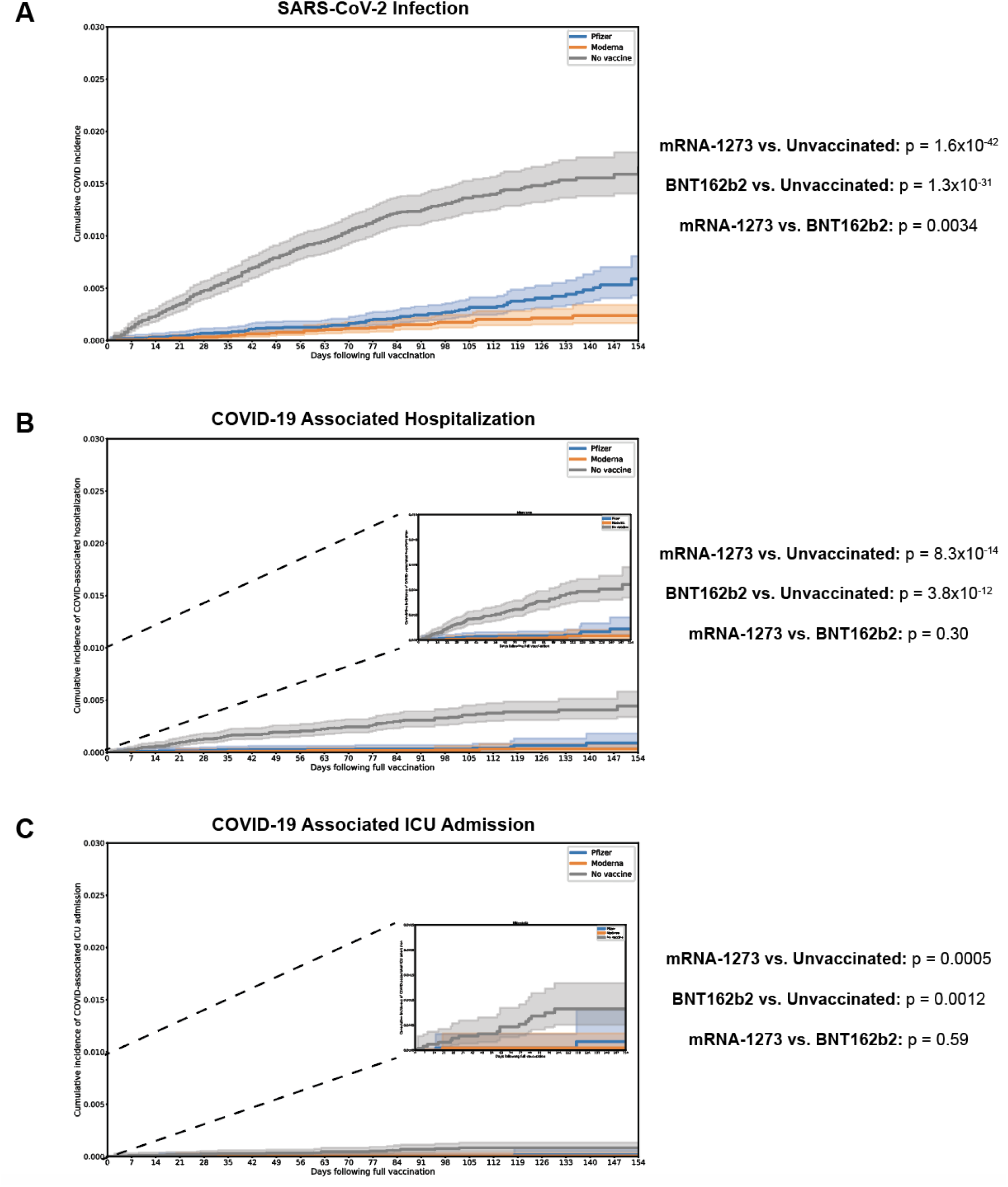
Kaplan-Meier analysis comparing the cumulative incidence of (A) SARS-CoV-2 infection, (B) COVID-19 associated hospitalization, and (C) COVID-19 associated ICU admission between the vaccinated and unvaccinated cohorts from Minnesota. Cumulative incidence at time *t* is the estimated proportion of individuals who experienced the outcome on or before time *t* (i.e., 1 minus the standard Kaplan-Meier survival estimate). This is assessed starting 14 days after the date of the actual or hypothetical second dose (i.e., starting on the date of full vaccination). The main figure in each panel has a y-axis ranging from 0 to 0.03. In (B) and (C), the inset plots are zoomed-in versions of the same plot with the y-axis ranging from 0 to 0.01. In each case, the log-rank p-value for each pairwise comparison is shown to the right of the plot.

**Figure S3.**
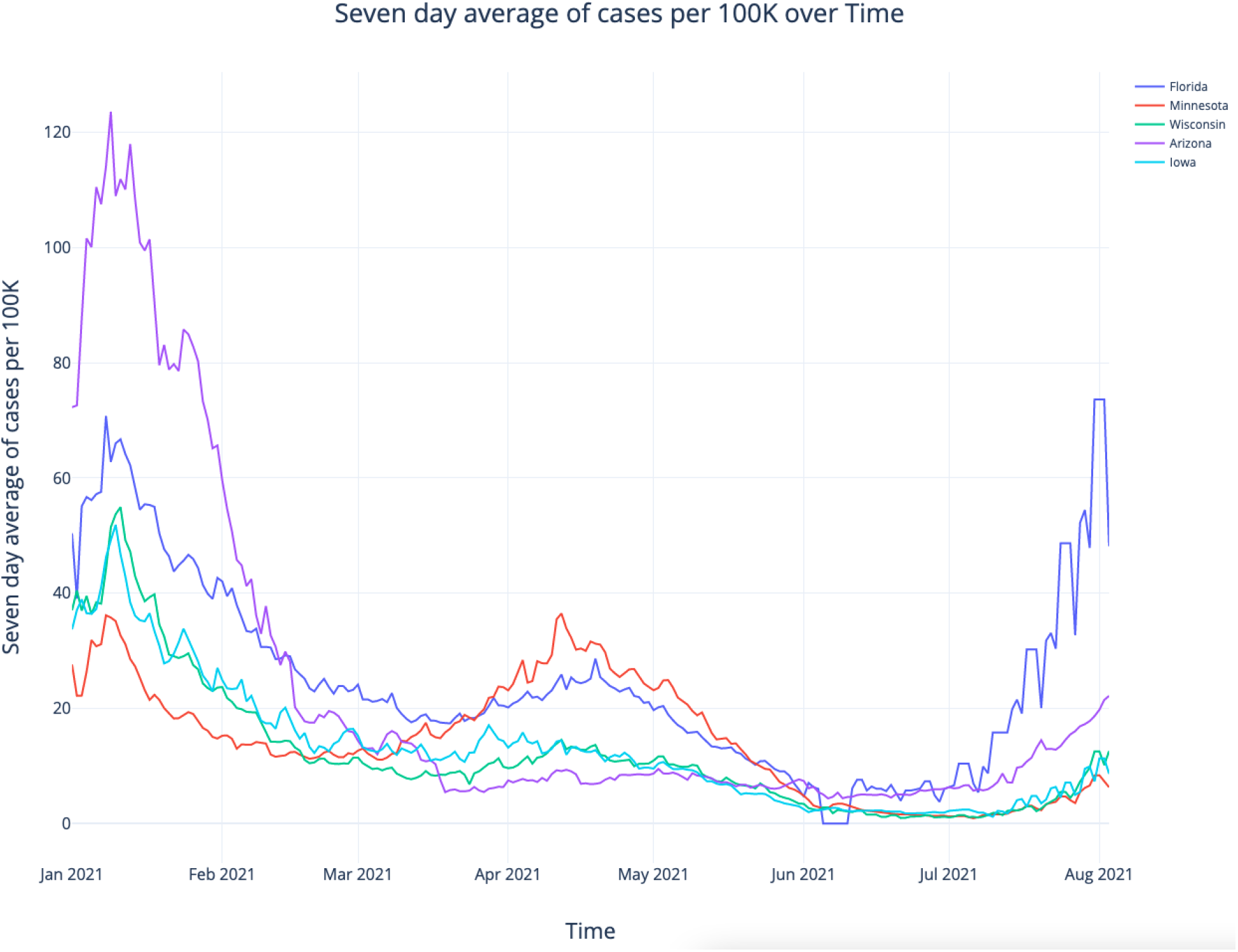
Number of COVID-19 cases per week in Florida, Minnesota, Wisconsin, Arizona and Iowa between January and July 2021. Data was accessed from New York Times.^23^

**Figure S4.**
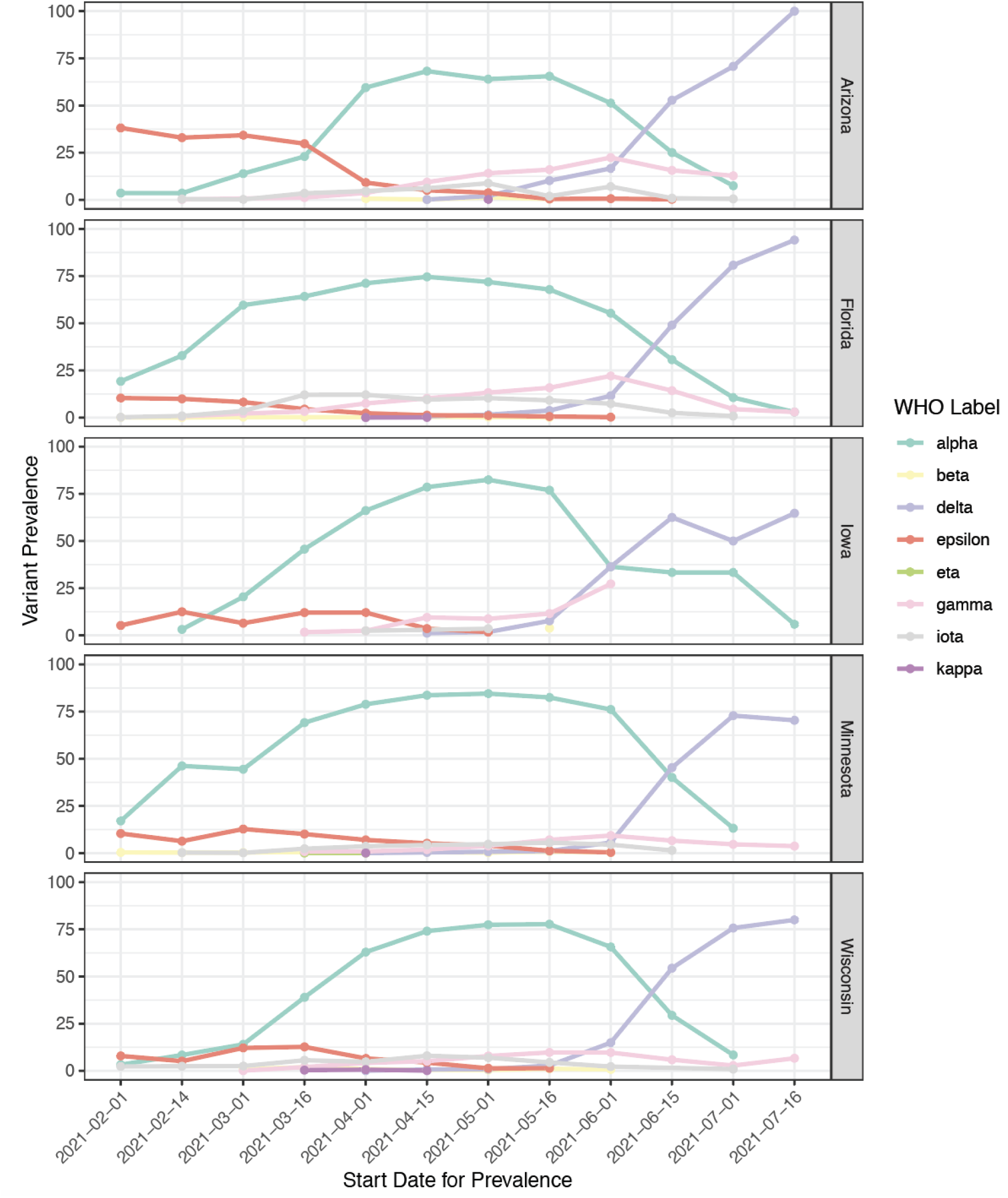
Prevalence of SARS-CoV-2 variants of concern and interest in US States with Mayo Clinic sites included in this analysis. Data is shown from February 2021 onward, the time period during which the effectiveness of full vaccination was assessed in this study. Data was accessed from the GISAID Initiative.^15^

**Figure S5.**
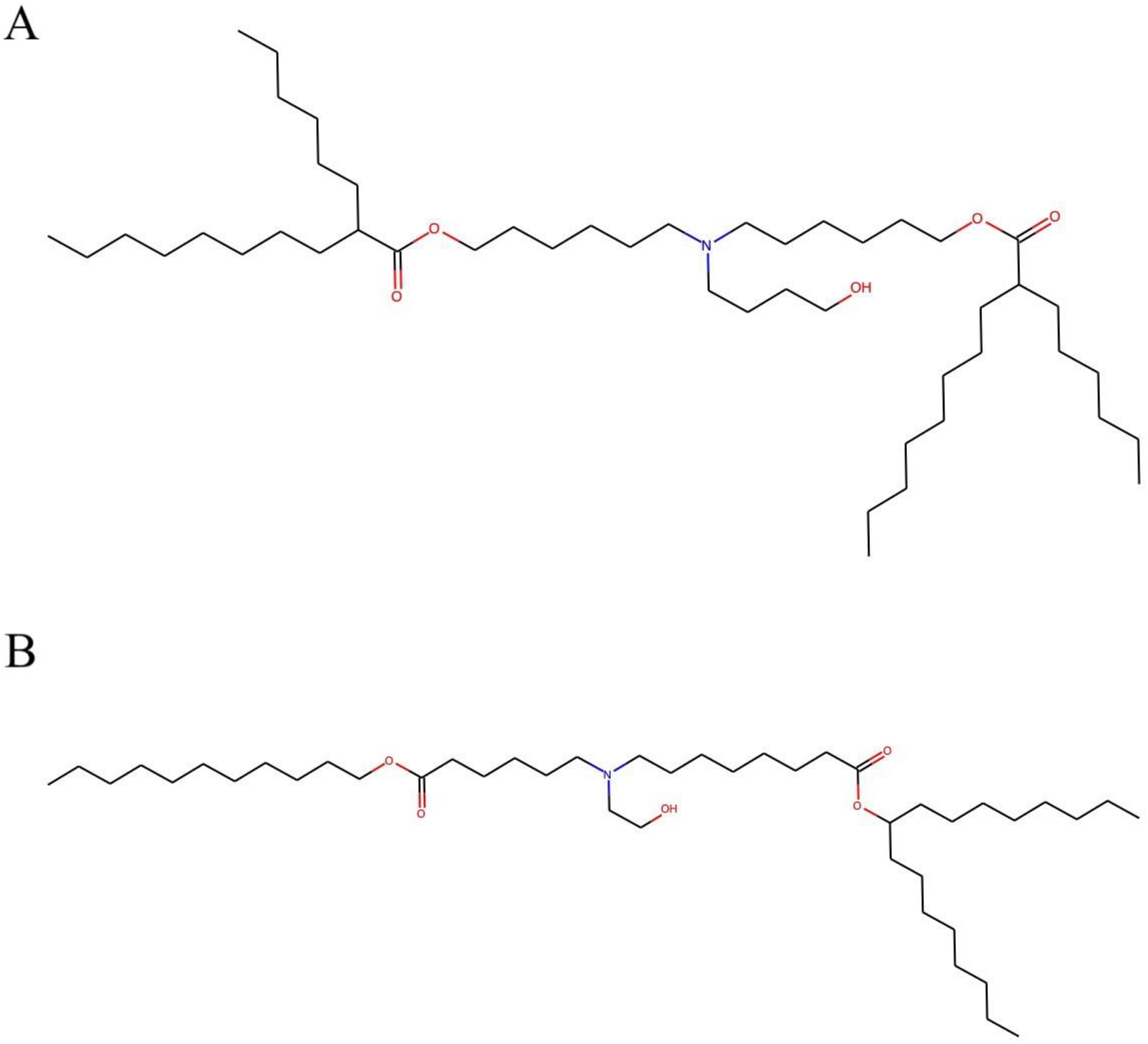
Comparison of the cationic lipid components of BNT162b2 and mRNA-1273. (A) ALC-0315 is the cationic lipid component of the BNT162b2 lipid nanoparticle. (B) SM-102 is the cationic lipid component of the mRNA-1273 lipid nanoparticle.

**Table S1.**
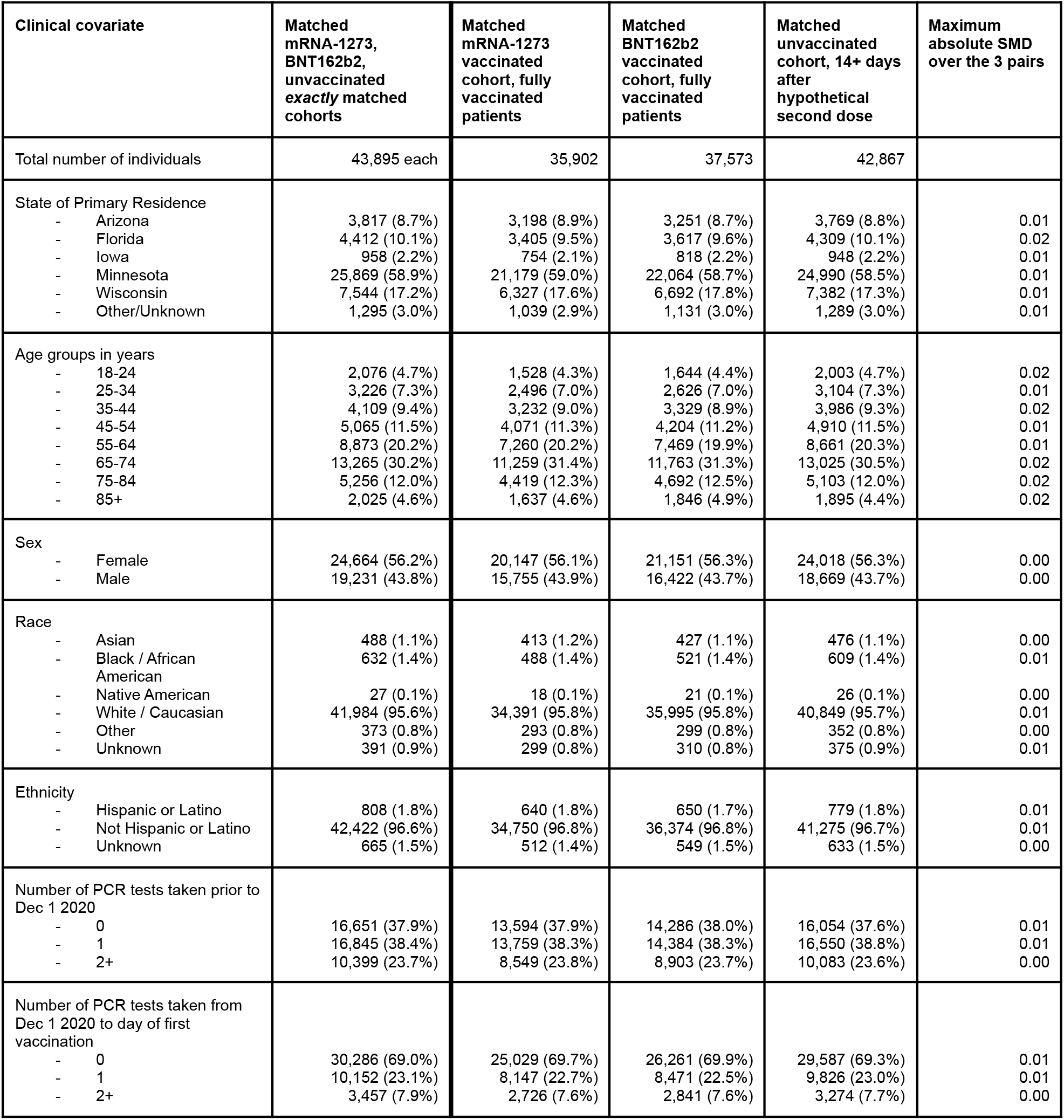
Clinical characteristics of 1-to-1 matched mRNA-1273-vaccinated, BNT162b2-vaccinated, and unvaccinated cohorts from all states. Covariates for matching include demographics (age, sex, race, ethnicity), state of location, date of vaccination, and number of prior SARS-CoV-2 PCR tests. Covariate statistics are also shown for the sub-cohorts of each cohort which are counted in the full vaccination effectiveness analysis (i.e., patients who have received two doses of the given vaccine and had at least 14 days of follow-up after the second dose or hypothetical second dose, without a positive PCR test at any date prior to 14 days following the second dose).

